# Sex-specific proteomic signatures improve cardiovascular risk prediction for the general population without cardiovascular disease or diabetes

**DOI:** 10.1101/2024.12.03.24318393

**Authors:** Ruijie Xie, Tomislav Vlaski, Sha Sha, Hermann Brenner, Ben Schöttker

**Affiliations:** Division of Clinical Epidemiology and Aging Research, German Cancer Research Center, Im Neuenheimer Feld 581, 69120 Heidelberg, Germany; Faculty of Medicine, Heidelberg University, 69115 Heidelberg, Germany

## Abstract

**Importance:** Accurate prediction of 10-year major adverse cardiovascular events (MACE) is crucial for effective cardiovascular disease prevention and management.

**Objective:** To evaluate whether adding sex-specific proteomic profiles to the SCORE2 model enhances 10-year MACE risk prediction in the large UK Biobank (UKB) cohort.

**Design, Setting, and Participants:** Data from 47,382 UKB participants, aged 40 to 69 years without prior cardiovascular disease or diabetes, were utilized. The cohort was randomly divided into derivation (70%) and validation (30%) sets.

**Exposures:** Proteomic profiling of plasma samples was conducted using the Olink Explore 3072 platform, measuring 2,923 unique proteins, of which 2,085 could be used. Sex-specific Least Absolute Shrinkage and Selection Operator (LASSO) regression was used for biomarker selection.

**Main Outcomes and Measures:** The primary outcome was 10-year MACE incidence, defined as cardiovascular death, non-fatal myocardial infarction, or non-fatal stroke. Model performance was assessed by changes in Harrell’s C-index, net reclassification index (NRI), and integrated discrimination index (IDI).

**Results:** During 10-year follow-up, 2,163 participants experienced MACE. Overall, 18 proteins were selected by LASSO regression, with 5 of them identified in both sexes, 7 only in males, and 6 only in females. Incorporating these proteins, significantly improved the C-index of the SCORE2 model from 0.713 to 0.778 (*P*□<□0.001) in the total population. The improvement was greater in males (C-index increase from 0.684 to 0.771; Δ□=□+0.087) than in females (from 0.720 to 0.769; Δ□=□+0.049). The NRI was 19.9% for the total population, 36.3% for males, and 18.2% for females. The WAP four-disulfide core domain protein (WFDC2), which modulates extracellular matrix degradation, impacting fibrosis and plaque stability, and the growth/differentiation factor 15 (GDF15), reflecting increased inflammatory activity, were the proteins contributing the strongest C-index increase in both sexes; even more than the N-terminal prohormone of brain natriuretic peptide (NTproBNP), which was also selected.

**Conclusions and Relevance:** The derived **s**ex-specific 10-year MACE risk prediction models, combining 12 protein concentrations among men and 11 protein concentrations among women with the SCORE2 model, significantly improved the discriminative abilities of the SCORE2 model. This study shows the potential of sex-specific proteomic profiles for enhanced cardiovascular risk stratification and personalized prevention strategies.

## Introduction

Cardiovascular disease (CVD) presents significant sex differences in both risk and underlying mechanisms.^1^ Men are generally at higher risk of developing CVD than women at any age.^2^ Women experience a notable increase in CVD risk later in life than men but often with more severe outcomes like heart failure with preserved ejection fraction and microvascular dysfunction.^3, 4^ Although these sex differences in CVD risk are well-established, our understanding of the underlying sex-specific biological mechanisms remains limited.^5^

The SCORE2 model introduced in 2021 represents a significant advancement in cardiovascular risk prediction by offering sex-specific competing risk algorithms.^6^ Derived from a comprehensive analysis of nearly 700,000 individuals across European cohorts, SCORE2 is designed to estimate the 10-year risk of major adverse cardiovascular events (MACE) in individuals without prior CVD or diabetes. Despite its clinical utility, further refinement is needed to enhance the precision of these risk assessments.^7, 8^

Proteomics enables comprehensive protein analysis, illuminating pivotal biological processes underlying CVD.^9^ This approach has the potential to uncover novel biomarkers that could improve our understanding of the complex interplay between genetic, environmental, and pathological factors in CVD progression.^10^ Despite evidence that proteomics can enhance cardiovascular risk prediction, previous studies often treat sex merely as a covariate and overlook the potential of sex-specific biomarker selection.^11–15^ Notably, research has identified widespread sex differences in circulating proteomic biomarkers, reflecting distinct pathways in the areas of inflammation, adiposity, and fibrosis that contribute to CVD development.^16^ Incorporating sex-specific protein concentration differences, mirroring these pathways, into predictive models may further improve the accuracy of cardiovascular risk assessment.

Thus, our study aimed to assess whether 10-year MACE risk prediction with the SCORE2 model can be improved by incorporating plasma protein concentrations, using a sex-specific selection approach. We used data from the large-scale UK Biobank (UKB) cohort, which recently released the proteomics data for 54,219 study participants for analysis by registered scientists.

## Methods

### Study population

The UKB is a large prospective cohort with 502,414 participants, aged 40 to 69 years, recruited from 13 March 2006 to 1 October 2010 across 22 assessment sites in England, Scotland, and Wales.^17^ The UKB was conducted in accordance with the Declaration of Helsinki. All participants provided written informed consent. A total of 54,219 participants have been selected for proteomics measurements, comprising a random subset of 46,595 individuals from the baseline UK Biobank cohort, 6,376 participants specifically selected to ensure adequate representation of 122 defined diseases, and 1,268 individuals who contributed to a COVID-19 research study. Therefore, the non-randomly selected participants are slightly (but not substantially) older and sicker than the randomly selected participants.^18^

**Supplemental Figure S1** shows the flowchart of exclusion criteria for the analyzed study population. The baseline blood samples of a total of 54,219 participants were selected for proteomics measurements. Participants with more than 50% missing values for measured proteins, with diabetes diagnoses or a history of MACE were excluded. Furthermore, participants with missing data on diabetes at baseline or MACE (either prior baseline or in the follow-up) were excluded. Finally, we included 47,382 individuals in the analysis. Overall, 89.2% of the included study participants were from the random subset. In the sensitivity analysis, the non-randomly selected participants (n=5,114) were excluded from the analyses.

### Plasma proteomics measurements

Proteomic profiling was performed on EDTA-plasma samples collected at baseline using the Olink Explore 3072 platform. More details can be found in **Appendix Text 1** (**Supplemental Material**). The platform detects 2,923 unique proteins. Proteins with more than 20% missing values or 25% of values below the limit of detection (N=838) were excluded. Ultimately, 2,085 proteins were included in the analysis.

### Variables of the SCORE2 model

The SCORE2 model, designed for adults without diabetes aged 40 to 69, includes age, sex, high-density lipoprotein cholesterol (HDL-C), total cholesterol, systolic blood pressure (SBP), and smoking status.^6^ Data on age, sex, and smoking status were obtained through standardized questionnaires. HDL-C and total cholesterol levels were measured using the Beckman Coulter AU5800 with an enzymatic method. SBP was recorded through automated readings using the Omron device on the left upper arm.

### Outcome ascertainment

The primary endpoint was MACE, defined as an endpoint of cardiovascular death, non-fatal myocardial infarctions, and non-fatal strokes, in alignment with the SCORE2 model.^6^ Occurrences of non-fatal myocardial infarctions and strokes were identified using primary care records or hospital episode statistics. Dates and causes of death were determined through death registries from the National Health Service Information Centre in England and Wales, and the National Health Service Central Register in Scotland. Participants were followed from baseline until the first occurrence of a MACE event, death, or the end of the ten-year follow-up period, whichever occurred first. A detailed definition of MACE is provided in **Supplemental Table S1**.

### Statistical analyses

#### General remarks

All analyses were performed using R software (version 4.3.0, R Foundation for Statistical Computing, Vienna, Austria). Statistical significance was defined as *P*-values < 0.05 for two-sided tests. Missing values of variables of the SCORE2 (variable with the highest proportion of missing values was HDL-C with 12.9%) and proteins (mostly complete with a few proteins with up to 20% of missing values) were single imputed using the chained equations method with random forest algorithms implemented in the R package *miceRanger* (version 1.5.0).

#### Biomarker Selection and Model Derivation

The UKB dataset was randomly split into a derivation set (70%) and a validation set (30%) for both male and female participants separately. Protein selection was conducted independently within each sex stratum to develop sex-specific risk algorithms.

Feature selection was performed using the Least Absolute Shrinkage and Selection Operator (LASSO) regression.^19^ A nested ten-fold cross-validation was conducted within 200 bootstrap samples. For each bootstrap sample, ten-fold cross-validation was used to optimize the regularization parameter λ by minimizing the validation error. Proteins were ranked based on their selection frequency across the 200 bootstrap iterations. We designated proteins selected in at least 95% of the bootstrap samples (score ≥ 190) as proteins of interest. This high threshold was chosen to enhance model generalizability and minimize overfitting, ensuring that only the most robust predictors were included.^20^ The selected proteins were then incorporated into the SCORE2 model to develop new sex-specific risk algorithms.

#### Model Performance Validation

The predictive performance of the derived model was validated using the remaining 30% of the UKB dataset as the validation set. Discrimination was assessed using Harrell’s C-index and the receiver operating characteristic (ROC) curve. The statistical significance of improvements in the C-index was evaluated using the method for comparing correlated C-indices in survival analysis proposed by Kang et al.,^21^ which is implemented in the R package *compareC* (version 1.3.2). Risk reclassification was evaluated using the net reclassification index (NRI) and the integrated discrimination index (IDI).^22^ Pre-specified cardiovascular risk categories (0–5%, >5–10%, and >10%) were applied to the NRI to determine the proportion of individuals correctly reclassified compared to the SCORE2 model. Model calibration was assessed by plotting observed MACE event rates against predicted rates across deciles of absolute predicted risk. Furthermore, the incremental contribution of each protein to discrimination was evaluated based on the increase in the C-index.

#### Associations of Selected Proteins with MACE

To report hazard ratios (HRs) and 95% confidence intervals (CIs) for the selected proteins (per one standard deviation increment) associated with 10-year MACE incidence in male and female participants, the proteins were individually added to Cox proportional hazards regression models in the validation set. These models were adjusted for SCORE2 model variables using the *survival* package (version 3.5-5) in R.

## Results

### Baseline characteristics and MACE case numbers

**Table 1** presents the baseline characteristics of the 47,382 UKB participants included in the analysis. The mean age was 56.4□±□8.2 years, and 44.1% were male. Over a 10-year follow-up period, 2,163 participants experienced MACE. Compared to participants who did not experience MACE, those who developed MACE were significantly older (mean age, 61.2□±□8.1 years vs. 56.2□±□8.2 years), were more frequently male (62.3% vs. 43.2%), had higher SBP (147.0□±□15.3 mmHg vs. 139.1□±□13.7 mmHg), lower HDL-C levels (1.4□±□0.4 mmol/L vs. 1.5□±□0.4 mmol/L), and were more frequently current smokers (18.9% vs. 10.0%).

**Table 1.**
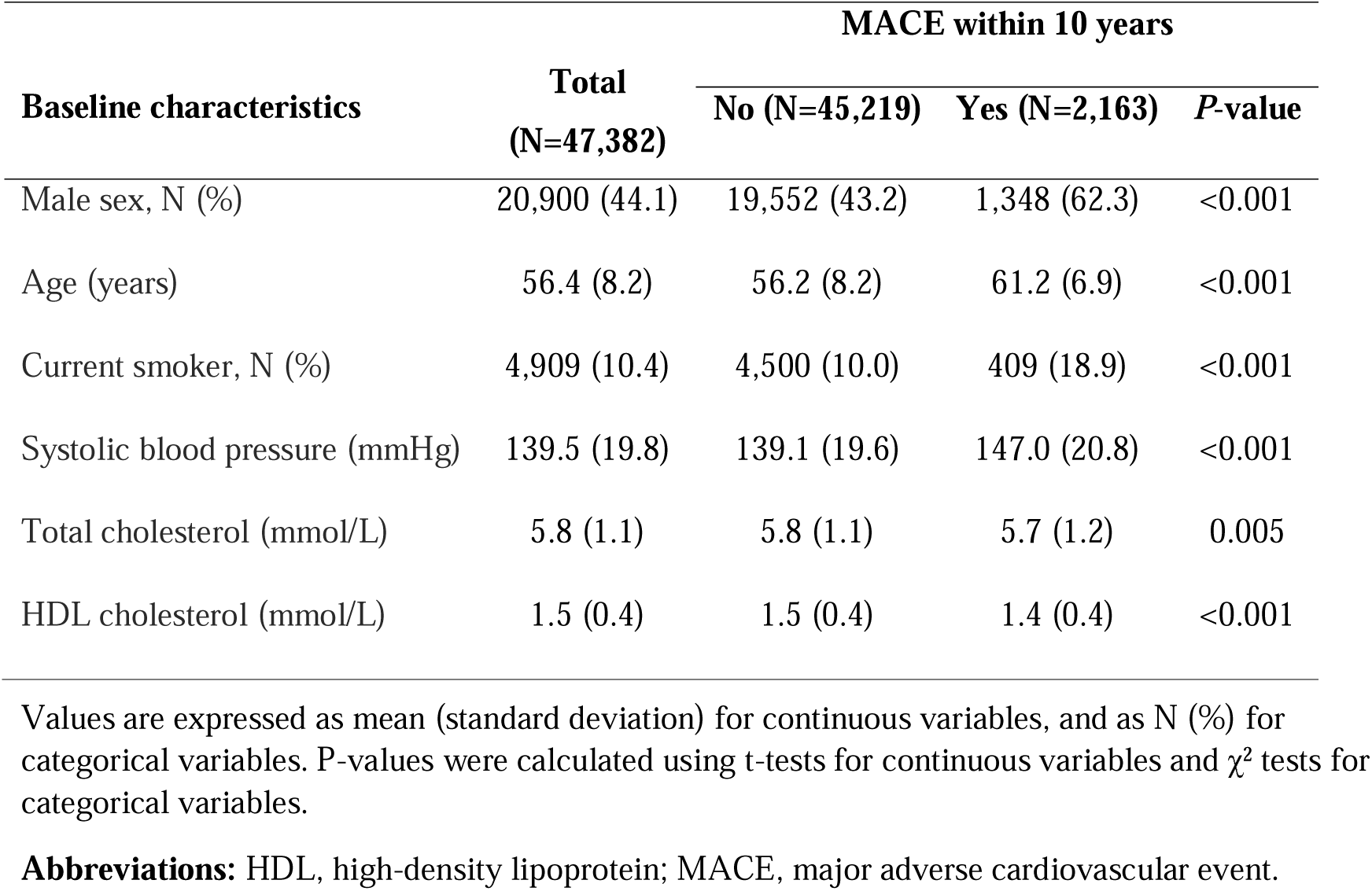
Baseline characteristics of selected participants from the UK Biobank.

| Baseline characteristics | Total<br>(N=47,382) | MACE within 10 years |  | P-value |
| --- | --- | --- | --- | --- |
|  |  | No (N=45,219) | Yes (N=2,163) |  |
| Male sex, N (%) | 20,900 (44.1) | 19,552 (43.2) | 1,348 (62.3) | <0.001 |
| Age (years) | 56.4 (8.2) | 56.2 (8.2) | 61.2 (6.9) | <0.001 |
| Current smoker, N (%) | 4,909 (10.4) | 4,500 (10.0) | 409 (18.9) | <0.001 |
| Systolic blood pressure (mmHg) | 139.5 (19.8) | 139.1 (19.6) | 147.0 (20.8) | <0.001 |
| Total cholesterol (mmol/L) | 5.8 (1.1) | 5.8 (1.1) | 5.7 (1.2) | 0.005 |
| HDL cholesterol (mmol/L) | 1.5 (0.4) | 1.5 (0.4) | 1.4 (0.4) | <0.001 |

Values are expressed as mean (standard deviation) for continuous variables, and as N (%) for categorical variables. P-values were calculated using t-tests for continuous variables and $\chi^2$ tests for categorical variables.
**Abbreviations:** HDL, high-density lipoprotein; MACE, major adverse cardiovascular event.

### Associations of selected proteins with MACE

A total of 18 proteins were selected to enhance MACE risk prediction in the SCORE2 model within the derivation set using LASSO analysis and bootstrapping. Among these, 5 proteins were identified in both sexes, 7 were specific to males, and 6 were specific to females (**Supplemental Table S2**).

Figure 1 illustrates the hazard ratios and confidence intervals for each of the selected proteins in males and females. Among the 18 proteins, 14 were significantly associated with MACE in both sexes. Among the remaining 4 proteins, ADAMTS13 (a disintegrin and metalloproteinase with thrombospondin motifs 13), BCAN (brevican core protein), and CXCL17 (C-C motif chemokine 7) showed significant associations with MACE in males but not in females, and CRYBB2 (Beta-crystallin B2) was significantly associated with MACE only in females.

**Figure 1.**
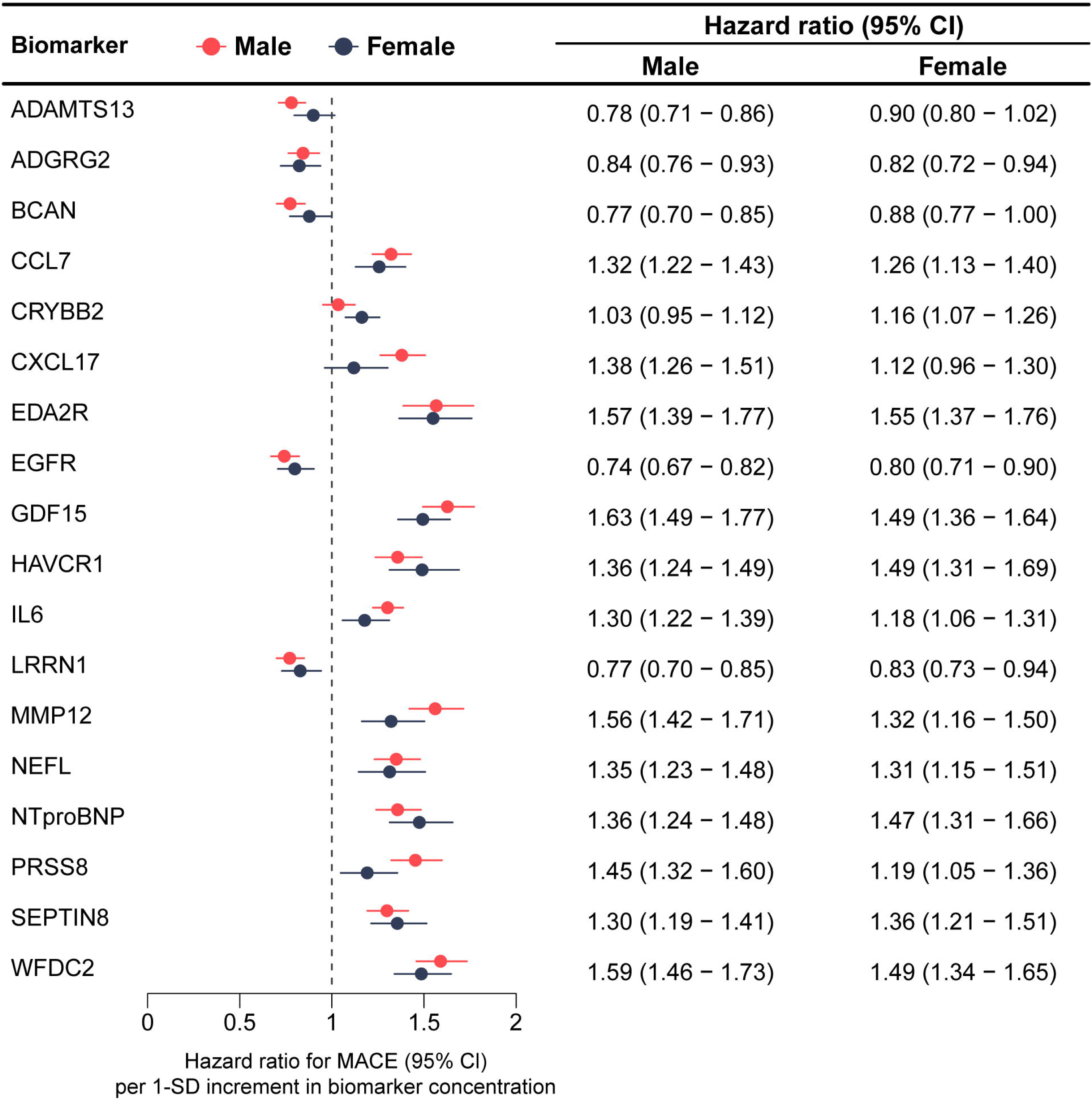
Associations between selected proteins and major cardiovascular events across sexes in the validation cohort (30% of UK Biobank, N=14,216) **Abbreviations:** ADAMTS13, A disintegrin and metalloproteinase with thrombospondin motifs 13; ADGRG2, Adhesion G-protein coupled receptor G2; BCAN, Brevican core protein; CCL7, C-C motif chemokine 7; CI, confidence interval; CRYBB2, Beta-crystallin B2; CXCL17, C-X-C motif chemokine 17; EDA2R, Tumor necrosis factor receptor superfamily member 27; EGFR, Epidermal growth factor receptor; GDF15, Growth/differentiation factor 15; HAVCR1, Hepatitis A virus cellular receptor 1; IL6, Interleukin-6; LRRN1, Leucine-rich repeat neuronal protein 1; MACE, major adverse cardiovascular events; MMP12, Macrophage metalloelastase; NEFL, Neurofilament light polypeptide; NPX, Normalized Protein eXpresstion; NTproBNP, N-terminal prohormone of brain natriuretic peptide; PRSS8, Prostasin; SEPTIN8, Septin-8; WFDC2, WAP four-disulfide core domain protein 2.

### MACE risk prediction by proteomic biomarkers

The ß-coefficients for all variables in the extended risk model are detailed in **Supplemental Table S2**. **Table 2** presents the predictive performance metrics of the SCORE2 model for 10-year MACE risk prediction, extended by the selected 18 proteins. In the derivation set, the integration of these proteins significantly improved the C-index of the SCORE2 model from 0.713 (95% CI: 0.701, 0.725) to 0.778 (95% CI: 0.767, 0.790) for the total population. This enhancement was observed in both sexes, with the C-index rising statistically significantly from 0.682 (95% CI: 0.666, 0.699) to 0.763 (95% CI: 0.747, 0.779) in males, and from 0.718 (95% CI: 0.699, 0.738) to 0.780 (95% CI: 0.751, 0.798) in females.

**Table 2.** Metrics of the predictive performance of the SCORE2 model for 10-year MACE risk without and with extension by proteins.

| Metrics | Male * | Female † | Overall |
| --- | --- | --- | --- |
| **Derivation set (70% of UK Biobank)** |  |  |  |
| **Total sample size (N=33,166) / MACE case number (N=1,492)** |  |  |  |
| C-Statistics (SCORE2) | 0.682 (0.666,0.699) | 0.718 (0.699,0.738) | 0.713 (0.701,0.725) |
| C-Statistics (+Proteins) | 0.763 (0.747,0.779) | 0.780 (0.751, 0.798) | 0.778 (0.767,0.790) |
| *P*-values for C-Statistics comparisons | **<0.001** | **<0.001** | **<0.001** |
| **Validation set (30% of UK Biobank)** |  |  |  |
| **Total sample size (N=14,216) / MACE case number (N=671)** |  |  |  |
| C-Statistics (SCORE2) | 0.684 (0.659, 0.705) | 0.720 (0.690, 0.751) | 0.716 (0.698, 0.734) |
| C-Statistics (+Proteins) | 0.771 (0.748,0.795) | 0.769 (0.740,0.798) | 0.778 (0.761,0.796) |
| *P*-values for C-Statistics comparisons | **<0.001** | **<0.001** | **<0.001** |
| NRI total (%) | **36.3 (25.3, 57.1)** | **18.2 (10.2, 26.5)** | **19.9 (14.7, 25.2)** |
| NRI events (%) | -3.6 (-9.2, 5.2) | **19.8 (12.6, 27.5)** | **13.4 (6.2, 17.2)** |
| NRI non-events (%) | **39.9 (26.3, 62.0)** | **-1.6 (-2.7, -0.7)** | **6.5 (4.0, 11.0)** |
| IDI | **0.058 (0.040, 0.075)** | **0.031 (0.016, 0.045)** | **0.045 (0.033, 0.058)** |

**Abbreviations:** IDI, integrated discrimination index; MACE, major cardiovascular events; NRI, net reclassification index.
\* Proteins included in the combined model for males were: ADAMTS13, BCAN, CCL7, CXCL17, GDF15, IL6, LRRN1, MMP12, NEFL, NTproBNP, PRSS8, WFDC2.
† Proteins included in the combined model for females were: ADGRG2, BCAN, CRYBB2, EDA2R, EGFR, GDF15, HAVCR1, MMP12, NTproBNP, SEPTIN8, WFDC2.

In the validation set, these findings were confirmed, showing similar improvements in the C-index with the C-index increasing significantly from 0.716 (0.698, 0.734) to 0.778 (0.761,0.796) in the total population, from 0.684 (0.659, 0.705) to 0.771 (0.748, 0.795) in males, and from 0.720 (0.690, 0.751) to 0.769 (0.740,0.798) in females. Figure 2 illustrates the ROC curves comparing the SCORE2 model with and without the inclusion of the proteomic data in the validation set, showing an improved discrimination performance with the addition of the proteins especially in males. In sensitivity analysis, excluding the non-randomly selected participants, the predictive performance of both the SCORE2 model and the proteins-extended model remained essentially unchanged in the total sample (**Supplemental Table S3**).

**Figure 2.**
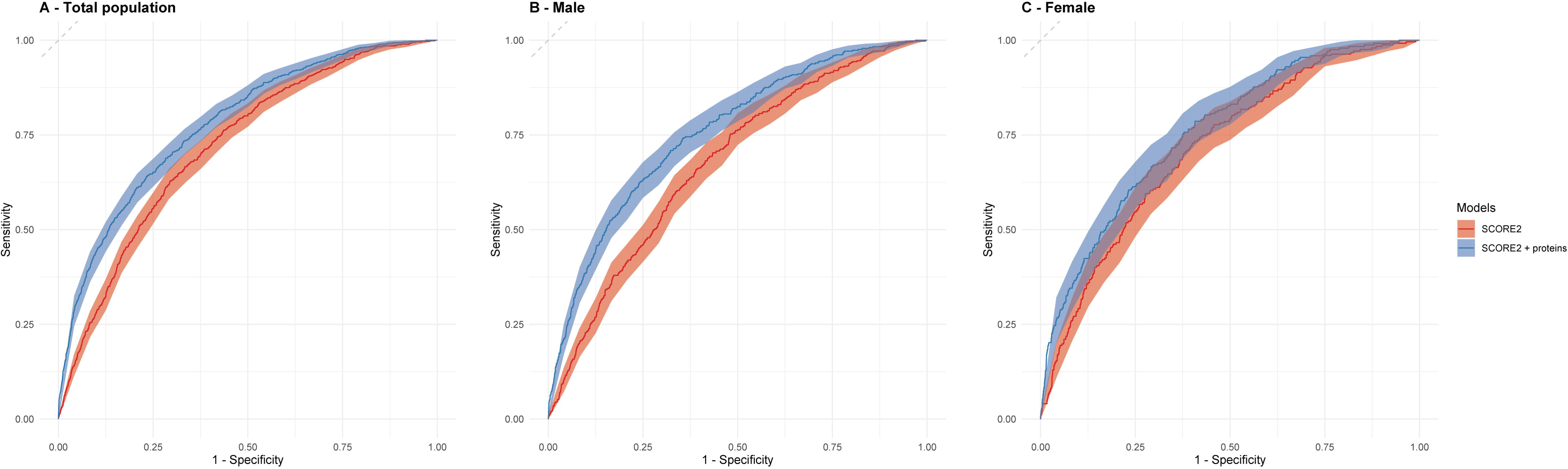
Receiver operating characteristic (ROC) curves for the SCORE2 model with and without proteomic biomarkers in the validation cohort (30% of UK Biobank, N=14,216) The ROC curves for the SCORE2 model (red line) and the SCORE2 model augmented with proteomic biomarkers (blue line) for predicting 10-year major adverse cardiovascular events (MACE). The results are shown for the total population (**Panel A**), males (**Panel B**), and females (**Panel C**).

Figure 3 illustrates the incremental improvement in the C-statistic achieved by incorporating each of the selected proteomic biomarkers. In males, all 12 selected proteins significantly improved the discriminative ability of the SCORE2 model. Notably, GDF15 (Growth/differentiation factor 15), WFDC2 (WAP four-disulfide core domain protein 2), MMP12 (Macrophage metalloelastase), CXCL17 (C-X-C motif chemokine 17), and IL6 (Interleukin-6) increased the C-index by more than 0.03 index points. In females, out of the 11 selected proteins, 6 significantly improved the SCORE2 model’s discriminative ability, while the remaining 5 proteins also contributed to model improvement, albeit without reaching statistical significance. This difference may be attributed to the already high baseline predictive ability of the SCORE2 model in females (0.720 compared to 0.684 in males). In agreement with the results observed in males, GDF15 and WFDC2 were the two most impactful biomarkers in females, with GDF15 increasing the C-index by 0.026 and WFDC2 by 0.035 index points.

**Figure 3.**
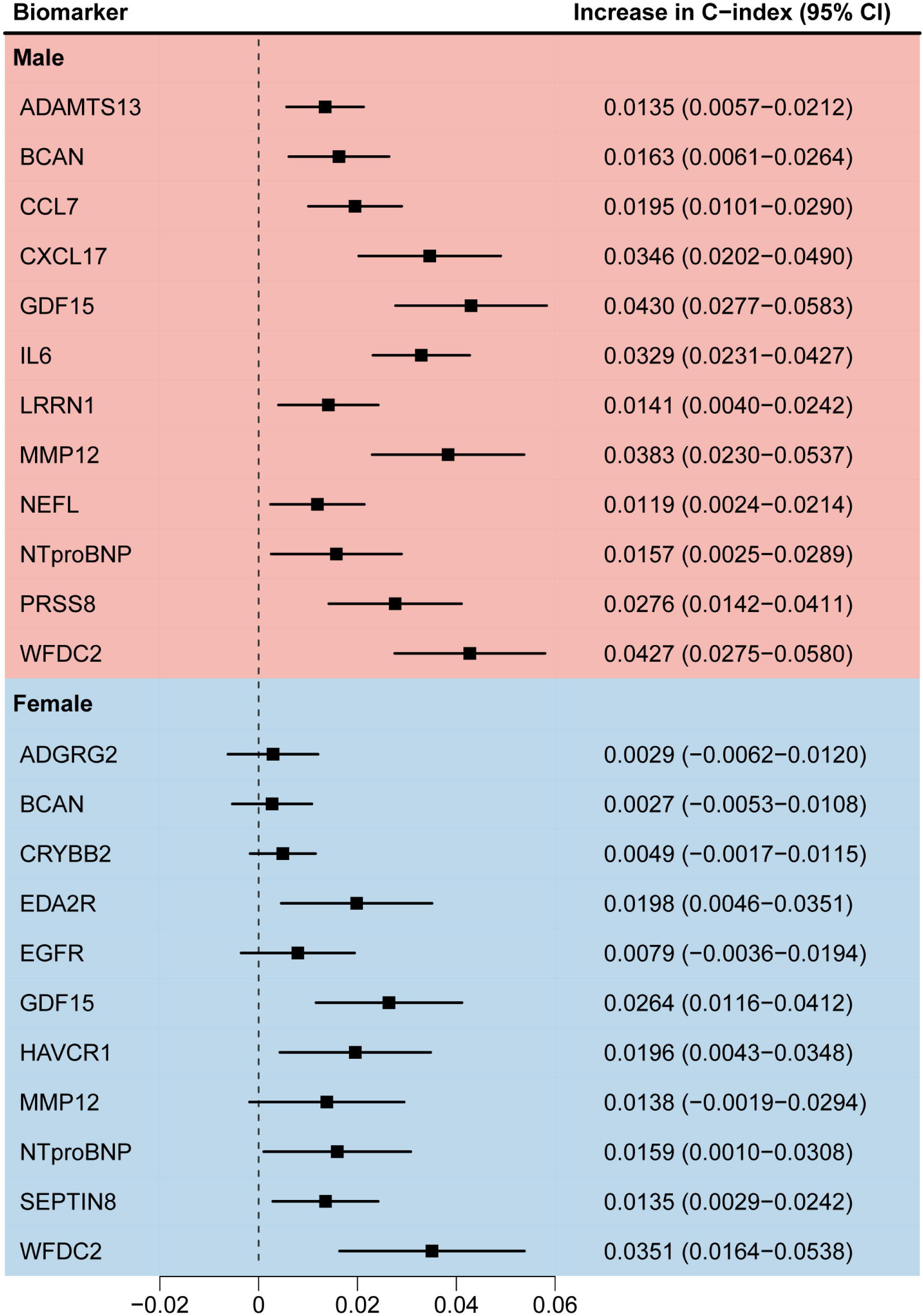
Contribution of individual proteomic biomarkers to the C-index in MACE risk prediction stratified by sex in the validation cohort (30% of UK Biobank, N=14,216) Contribution of each selected proteomic biomarker to the improvement in the C-index, stratified by sex, for predicting 10-year MACE. **Abbreviations:** see legend of Figure 1.

Additionally, adding the proteomic signatures to the SCORE2 model demonstrated significant gains in reclassification statistics, with a notable increase in the NRI for the total population, reaching 19.9% (95% CI: 14.7, 25.2). The NRI was particularly high in males with 36.3% (95% CI: 25.3, 57.1) but the NRI in females was notable as well (18.2% (95% CI: 10.2, 26.5)). In males, more non-events were correctly re-classified than events. In females, it was the opposite, with more events correctly re-classified than non-events. Taken together in the total population, the correct re-classification outweighs the incorrect re-classifications in both events and non-events. This is supported by statistically significant IDIs for the total population, males, and females. **Supplemental Figure S2** shows the re-classification in more detail, with re-classification tables for low-risk (≤5%), intermediate risk (5-10%) and high-risk (>10%) categories with respect to MACE.

Calibration curves of both the original and extended SCORE2 models in the internal validation set are depicted in **Supplemental Figure S3.** Both are very well calibrated.

## Discussion

### Summary of the Findings

Our study demonstrates that incorporating sex-specific proteomic biomarkers into the SCORE2 model significantly improves the prediction of 10-year MACE risk in individuals without prior cardiovascular disease or diabetes. Overall, 18 proteins were selected, with 5 of them identified in both sexes, 7 only in males, and 6 only in females. WFDC2 and GDF15 contributed the highest increase in C-index in both sexes. Furthermore, high NRIs were observed for both sexes, with a particularly high NRI of 36.3% for males.

### Comparison with Previous Studies

Several prior studies have demonstrated the potential of proteomic biomarkers to enhance cardiovascular risk prediction.^11–15, 23–26^ For instance, in the EPIC-Norfolk cohort, incorporating 50 proteins into a cardiovascular risk model resulted in an AUC improvement of 0.024 for predicting myocardial infarction and 0.071 for predicting cardiovascular events over three years.^11^ Similarly, in an Icelandic cohort, an increase in the C-index by 0.014 was observed for predicting MACE when a protein risk score was added to a traditional risk model.^12^

Additionally, the study of Royer et al. investigated the enhancement of SCORE2’s predictive capability for MACE with proteomic biomarkers in the same data set we used from the UK Biobank.^14^ The main difference to our analysis is that no sex-specific selection procedure was used. Royer et al. reported that integrating 114 proteins into the SCORE2 model increased the AUC by 0.031 (from 0.740 to 0.771) and achieved an NRI of 14.0%.^14^ Our sex-specific selection procedure achieved an exactly twice as high improvement of model discrimination (C-index increase of 0.062) from 0.716 to 0.778) and the C-index of the final combined model in our analysis (0.778) was comparable to that of the model of Royer et al. (0.771). However, we achieved this C-index with adding only 12 proteins for males and 11 proteins for females to the SCORE2 model instead of 114 proteins. In addition, the NRI was higher in our analysis (19.9%) than in the one of Royer et al. (14.0%). This notable enhancement in the NRI with only a few proteins highlights the effectiveness of a sex-specific approach in improving predictive accuracy. By accounting for biological differences and a different underlying baseline cardiovascular risk of males and females, our model more precisely identifies individuals at high risk for MACE in the next 10 years who could benefit from targeted preventive strategies. Finally, it is more cost-effective and easier to translate into clinical routine to measure up to 12 proteins than measuring 114 proteins.

### Biological Mechanisms of the 18 Selected Proteins for MACE Risk Prediction

GDF15 and WFDC2 increased the discriminative ability of the model for MACE prediction in both sexes most. Elevated levels of GDF15 reflect heightened inflammatory activity and are predictive of adverse cardiovascular outcomes.^27^ With IL6, CCL7, CXCL17, and the tumor necrosis factor receptor superfamily member 27 (EDA2R) further proteins involved in inflammation were selected. IL6 is a pro-inflammatory cytokine that promotes endothelial dysfunction and plaque formation, leading to increased cardiovascular risk.^28^ CCL7 recruits immune cells to inflammatory sites, contributing to plaque instability.^29^ CXCL17 is involved in immune cell chemotaxis and may accelerate atherogenesis.^30^ EDA2R mediates TNF-related cytokine signaling, exacerbating vascular inflammation and plaque vulnerability.^31^

WFDC2 modulates extracellular matrix degradation, impacting fibrosis and plaque stability.^32^ Further proteins selected for the MACE risk algorithms, which are involved in extracellular matrix remodeling, were MMP12, ADAMTS13, and PRSS8 (Prostasin). MMP12 degrades extracellular components, weakening the fibrous cap, which increases rupture risk.^33^

ADAMTS13 regulates the von Willebrand factor cleavage, affecting thrombosis risk.^34^ PRSS8 influences vascular stiffness and hypertension through sodium balance regulation.^35^

A third large group of selected proteins affect vascular function through neural pathways. These were BCAN, NEFL (Neurofilament light polypeptide), LRRN1 (Leucine-rich repeat neuronal protein 1), CRYBB2, and SEPTIN8 (Septin-8). BCAN may influence vascular integrity via extracellular matrix interactions.^36^ NEFL reflects neurodegenerative processes impacting autonomic cardiovascular regulation.^37^ LRRN1 is involved in neuronal signaling affecting vascular tone.^38^ CRYBB2 may be involved in oxidative stress pathways or hormonal regulation affecting cardiovascular health in women.^39^ SEPTIN8 is involved in cytoskeletal organization and contributes to atherosclerosis by influencing cell migration and proliferation within the vessel wall.^40^

The remaining selected proteins have rather specific functions. NTproBNP (N-terminal prohormone of brain natriuretic peptide) is a well-established biomarker reflecting cardiac wall stress and dysfunction.^41^ EGFR (Epidermal growth factor receptor) plays a role in vascular smooth muscle cell proliferation and neointimal formation, and its activation may lead to vascular remodeling and plaque progression.^42^ ADGRG2 (Adhesion G-protein coupled receptor G2) is involved in cell adhesion and signaling, potentially affecting endothelial function and vascular integrity.^43^ HAVCR1 (Hepatitis A virus cellular receptor 1) is implicated in immune regulation and may contribute to atherosclerosis by modulating immune cell activity and endothelial interactions.^44^

### Strengths and Limitations

The strengths of this study include its large sample size and the comprehensive evaluation of a wide array of proteomic biomarkers. The sex-specific biomarker selection led to better risk prediction models and supports a personalized risk assessment. By focusing on proteins with high selection frequency in bootstrap samples, we minimized overfitting and improved model generalizability.

However, there are limitations to consider. We did not test the robustness of our model in external validation cohorts, which should be done by future research. Furthermore, future studies should calibrate and validate the model in other populations because the findings can currently only be generalized to the population of the UK and other Western European countries with low cardiovascular risk.

## Conclusion

The derived sex-specific 10-year MACE risk prediction models, adding the concentrations of 12 proteins for men and 11 proteins for women, significantly enhanced the discriminative abilities of the SCORE2 model for adults from the general population without diabetes mellitus included in the UK Biobank. Provided the model’s performance is similar in future external validation studies, it could be used to further improve cardiovascular prevention measures, which currently use the SCORE2 risk charts.

## Data Availability

Data from the UK Biobank are available to bona fide researchers upon application through the UK Biobank Access Management System (https://www.ukbiobank.ac.uk/enable-your-research/apply-for-access).

## ARTICLE INFORMATION

## Acknowledgements

We would like to thank all participants of the UK Biobank as well as the staff of the UK Biobank assessment centers for their contributions. This UK Biobank analysis was conducted under application no. 101633.

## Supplemental Materials

Appendix Text 1

Supplemental Tables S1–S2

Supplemental Figures S1–S3

## Non-standard Abbreviations and Acronyms

ADAMTS13: A disintegrin and metalloproteinase with thrombospondin motifs 13
ADGRG2: Adhesion G-protein coupled receptor G2
BCAN: Brevican core protein
CCL7: C-C motif chemokine 7
CI: Confidence intervals
CRYBB2: Beta-crystallin B2
CVD: Cardiovascular disease
CXCL17: C-X-C motif chemokine 17
EDA2R: Tumor necrosis factor receptor superfamily member 27
EGFR: Epidermal growth factor receptor
GDF15: Growth/differentiation factor 15
HAVCR1: Hepatitis A virus cellular receptor 1
HDL-C: High-density lipoprotein cholesterol
HR: Hazard ratio
IDI: Integrated discrimination index
IL6: Interleukin-6
LASSO: Least absolute shrinkage and selection operator
LRRN1: Leucine-rich repeat neuronal protein 1
MACE: Major adverse cardiovascular events
MMP12: Macrophage metalloelastase
NEFL: Neurofilament light polypeptide
NPX: Normalized Protein eXpression
NRI: Net reclassification index
NTproBNP: N-terminal prohormone of brain natriuretic peptide
PEA: Proximity Extension Assay
PRSS8: Prostasin
SBP: Systolic blood pressure
SD: Standard deviation
SEPTIN8: Septin-8
UKB: UK Biobank
WFDC2: WAP four-disulfide core domain protein 2

