## Supplemental Tables and Figures for "Sex-specific proteomic signatures improve cardiovascular risk prediction for the general population without cardiovascular disease or diabetes"

**Appendix Text 1 – Details of proteomics measurement**

Samples were shipped on dry ice to the Olink Analysis Service in Uppsala, Sweden, where the proteomic analyses were performed. The assay protocols, including sample handling and selection procedures, have been described previously.^1^ In brief, Olink utilizes the Proximity Extension Assay (PEA) method that targets proteins via pairs of antibodies linked to complementary oligonucleotides.^2-4^ Upon binding to specific proteins, these antibodies facilitate probe hybridization, amplification, and subsequent quantification via next-generation sequencing. Each protein-targeting assay on every plate establishes detection limits, determined from triplicate negative controls. Normalized protein expression (NPX) values are calculated by normalizing to the extension control, log2 transformation, and further normalization against plate controls. Samples are flagged if NPX values deviate by more than ±0.3 NPX from the plate median within an abundance block, or if the average assay count per sample drops below 500. Assays receive alerts if triplicate negative controls show a median deviation exceeding five standard deviations (SD) from preset values set by Olink.

**Supplemental Table S1.** Definition of endpoint major cardiovascular event (MACE)

| **Fatal MACE – cause-specific mortality due to any of the following:** | |
| --- | --- |
| *Endpoints included* | *ICD10-codes* |
| Hypertensive disease | I10-16 |
| Ischemic heart disease | I20-25 |
| Arrhythmias, heart failure | I46-52 |
| Cerebrovascular disease | I60-69 |
| Atherosclerosis/aortic aneurysm | I70-73 |
| Sudden death and death within 24 hours of symptom onset | R96.0-96.1 |
| *Endpoints excluded from the above endpoint:* | *ICD10-codes* |
| Myocarditis, unspecified | I51.4 |
| Subarachnoid haemorrhage | I60 |
| Subdural hemorrhage | I62 |
| Cerebral aneurysm | I67.1 |
| Cerebral arteritis | I68.2 |
| Moyamoya | I67.5 |
| **Non-fatal MACE** | *ICD10-codes* |
| Non-fatal myocardial infarction | I21-I23 |
| Non-fatal stroke | I61, I63-I66, I69 |

### **Supplemental** **Table S2.** ß-coefficients of the variables of the SCORE2 model extended by 18 proteins for 10-year prediction of major cardiovascular events

| **Risk factor (units)** | **Coefficient** | |
| --- | --- | --- |
|  | **Male** | **Female** |
| **SCORE2 variables** |  |  |
| Age (per 5 years) | 0.1882 | 0.2833 |
| Current smoking | 0.1601 | 0.4543 |
| Systolic blood pressure (per 20mmHg) | 0.1157 | 0.2152 |
| Total cholesterol (per 1 mmol/L) | 0.0772 | 0.0027 |
| HDL cholesterol (per 0.5 mmol/L) | -0.1286 | -0.0466 |
| Smoking interaction with age | -0.0378 | -0.0678 |
| SBP interaction with age | -0.0839 | -0.0464 |
| Total cholesterol interaction with age | -0.0351 | -0.0242 |
| HDL interaction with age | -0.0213 | 0.0326 |
| **Additional proteins** **(per 1 SD)** |  |  |
| ADAMTS13 | -0.5045 | - |
| ADGRG2 | - | -0.5115 |
| BCAN | -0.4317 | -0.3065 |
| CCL7 | 0.1692 | - |
| CRYBB2 | - | 0.1065 |
| CXCL17 | 0.1976 | - |
| EDA2R | - | 0.0039 |
| EGFR | - | -0.5132 |
| GDF15 | 0.2204 | 0.3188 |
| HAVCR1 | - | 0.1832 |
| IL6 | 0.0826 | - |
| LRRN1 | -0.1956 | - |
| MMP12 | 0.2937 | 0.2286 |
| NEFL | 0.2009 | - |
| NTproBNP | 0.1906 | 0.1801 |
| PRSS8 | 0.2465 | - |
| SEPTIN8 | - | 0.3420 |
| WFDC2 | -0.0738 | 0.2344 |

**Abbreviations:** ADAMTS13, A disintegrin and metalloproteinase with thrombospondin motifs 13; ADGRG2, Adhesion G-protein coupled receptor G2; BCAN, Brevican core protein; CCL7, C-C motif chemokine 7; CRYBB2, Beta-crystallin B2; CXCL17, C-X-C motif chemokine 17; EDA2R, Tumor necrosis factor receptor superfamily member 27; EGFR, Epidermal growth factor receptor; GDF15, Growth/differentiation factor 15; HAVCR1, Hepatitis A virus cellular receptor 1; IL6, Interleukin-6; LRRN1, Leucine-rich repeat neuronal protein 1; MMP12, Macrophage metalloelastase; NEFL, Neurofilament light polypeptide; NTproBNP, N-terminal prohormone of brain natriuretic peptide; PRSS8, Prostasin; SD, standard deviation; SEPTIN8, Septin-8; WFDC2, WAP four-disulfide core domain protein.

**Supplemental Table S3.** Sensitivity Analysis: Metrics of the predictive performance of the (protein extended) SCORE2 model for 10-year MACE risk including only randomly selected participants

| **Metrics** | **Male** **^*^** | **Female** **^†^** | **Overall** |
| --- | --- | --- | --- |
| **Derivation set (70% of UK Biobank)** | | | |
| **Total sample size (N=29,585) / MACE case number (N=1,164)** | | | |
| C-Statistics (SCORE2) | 0.676 (0.658,0.694) | 0.726 (0.704,0.749) | 0.727 (0.713,0.740) |
| C-Statistics (+Proteins) | 0.753 (0.735,0.770) | 0.774 (0.753, 0.794) | 0.780 (0.767,0.793) |
| *P*-values for C-Statistics comparisons | **<0.001** | **<0.001** | **<0.001** |
| **Validation set (30% of UK Biobank)** | | | |
| **Total sample size (N=12,683) / MACE case number (N=515)** | | | |
| C-Statistics (SCORE2) | 0.677 (0.649, 0.705) | 0.713 (0.678, 0.769) | 0.724 (0.703, 0.745) |
| C-Statistics (+Proteins) | 0.756 (0.729,0.783) | 0.747 (0.712,0.787) | 0.772 (0.752,0.792) |
| *P*-values for C-Statistics comparisons | **<0.001** | **<0.001** | **<0.001** |
| NRI total (%) | **22.1 (15.3, 29.2)** | **16.2 (8.7, 22.6)** | **20.2 (14.7, 24.7)** |
| NRI events (%) | 14.2 (-0.1, 22.6) | **19.5 (12.0, 26.7)** | **22.1 (16.5, 26.7)** |
| NRI non-events (%) | **7.0 (1.4, 21.7)** | **-3.3 (-4.2, -2.6)** | **-2.0 (-2.8, -1.0)** |
| IDI | **0.032 (0.022, 0.048)** | **0.015 (0.006, 0.028)** | **0.025 (0.018, 0.034)** |

**Abbreviations:** IDI, integrated discrimination index; MACE, major cardiovascular events; NRI, net reclassification index.

^*^ Proteins included in the combined model for males were: ADAMTS13, BCAN, CCL7, CXCL17, GDF15, IL6, LRRN1, MMP12, NEFL, NTproBNP, PRSS8, WFDC2.

^†^ Proteins included in the combined model for females were: ADGRG2, BCAN, CRYBB2, EDA2R, EGFR, GDF15, HAVCR1, MMP12, NTproBNP, SEPTIN8, WFDC2.

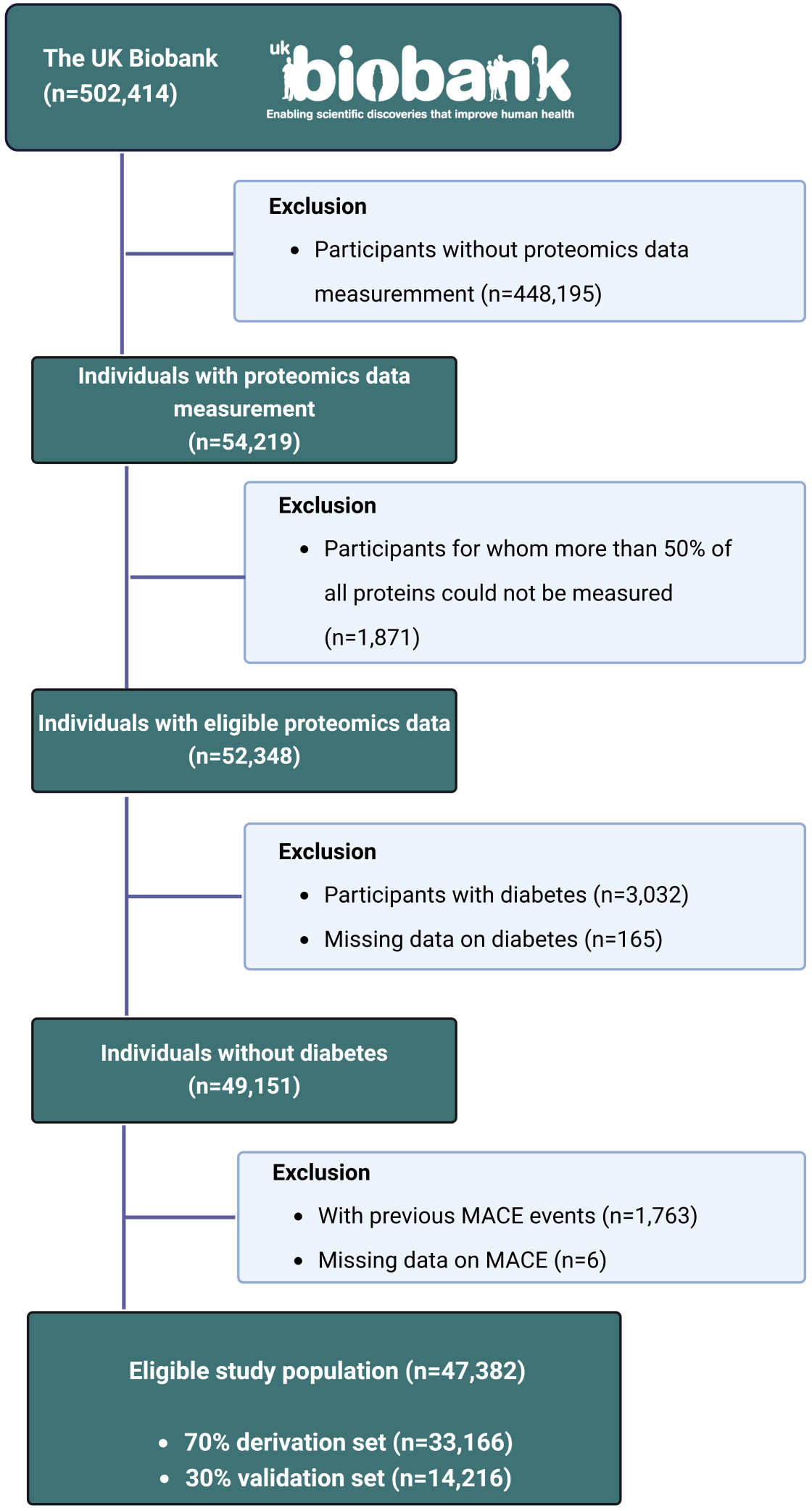

### **Supplemental** **Figure S1.** Flow-charts for participant inclusion and exclusion

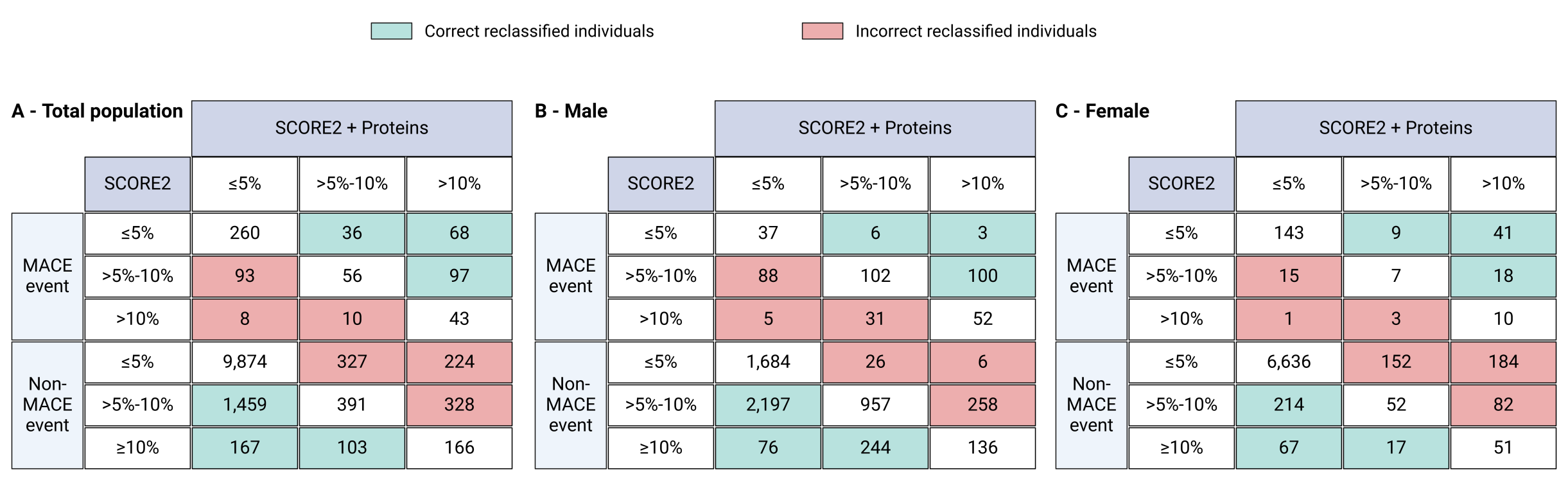

### **Supplemental Figure S2.** Reclassification of the SCORE2 model with and without proteomic biomarkers in the validation set (30% of UK Biobank, N=14,216)

The reclassification of individuals into cardiovascular risk categories (≤5%, >5%-10%, >10%) for major adverse cardiovascular events (MACE) using the SCORE2 model alone and with the addition of proteomic biomarkers. The results are shown for the total population (**Panel A**), males (**Panel B**), and females (**Panel C**).

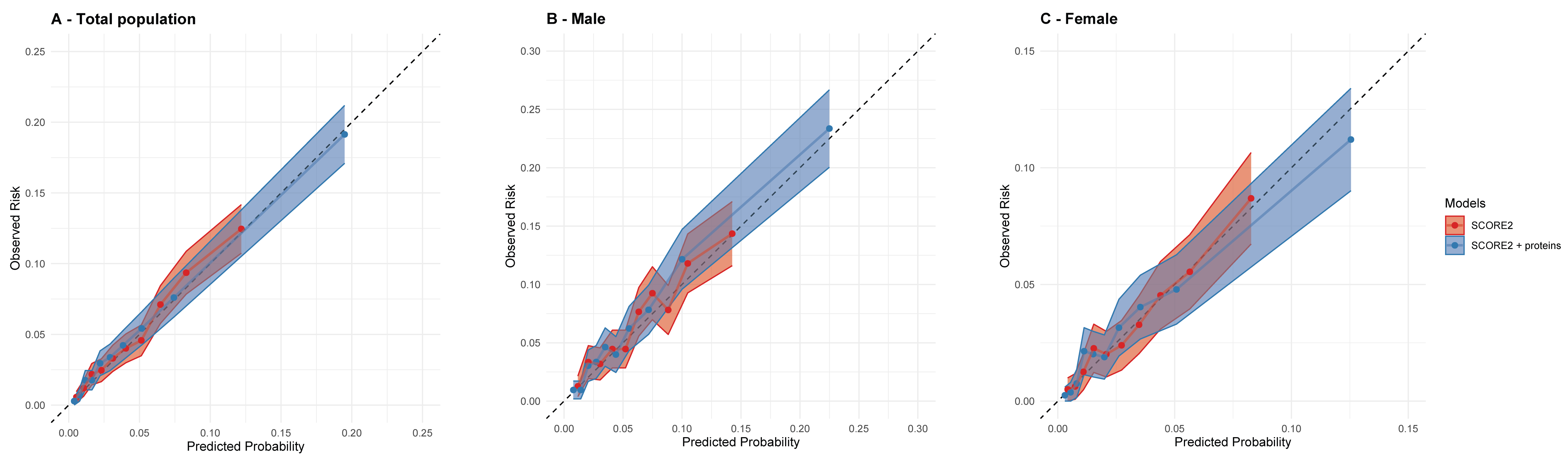

### **Supplemental Figure S3.** Calibration curves of the SCORE2 model with and without proteomics data for 10-year MACE risk prediction in the validation set (30% of UK Biobank, N=14,216)

Proteins that were included in the clinical CDRS were detailed in **Supplemental Table S2**.
